# Health Care Workers’ Mental Health During the First Weeks of the SARS-CoV-2 Pandemic in Switzerland – A Cross-Sectional Study

**DOI:** 10.1101/2020.05.04.20088625

**Authors:** Sonja Weilenmann, Jutta Ernst, Heidi Petry, Monique C. Pfaltz, Onur Sazpinar, Samuel Gehrke, Francesca Paolercio, Roland von Känel, Tobias R. Spiller

## Abstract

**Background:** The current SARS-CoV-2 pandemic poses various challenges for health care workers (HCWs), which may impair their mental health. First evidence from China suggests that HCWs are at risk for anxiety and depression. However, generalizability to western countries is limited. The current study aimed at exploring HCWs’ mental health during the SARS-CoV-2 pandemic in Switzerland. In addition, we conducted a network analysis to investigate the independent effect of risk and protective factors on HCWs’ mental health and their interplay.

**Methods:** In an exploratory, cross-sectional, nation-wide online survey, we assessed demographics, work characteristics, COVID-19 exposure, and anxiety, depression, and burnout in 857 physicians and 553 nurses during the pandemic in Switzerland. At the time of data collection, Switzerland had among the highest per capita rate of COVID-19 cases in the world.

**Results:** Overall symptom levels of anxiety, depression, and burnout were elevated. Women, nurses, frontline staff and HCWs exposed to COVID-19 patients reported more symptoms than their peers. However, these effects were all small and, in the network analysis, most of them did not remain significant after controlling for the other factors. Whereas COVID-19 exposure was only partially associated with mental health, perceived support by the employer independently predicted anxiety and burnout.

**Conclusions:** Our finding that HCWs had elevated levels of anxiety, depression, and burnout underscores the importance to systematically monitor HCWs’ mental health during this ongoing pandemic. Because perceived support and mental health impairments were negatively related, we encourage the implementation of supportive measures for HCWs’ well-being during this crisis.

## Introduction

Since December 2019, the world has witnessed a pandemic spread of SARS-CoV-2 with daily increasing numbers of patients suffering from COVID-19.^1,2^ This global public health crisis poses various challenges for health care workers (HCW) all around the world. Over the last weeks, some HCWs worked additional hours to care for the high number of COVID-19 patients and put themselves at risk for infection, while others have seen their workload diminish due to public health-related measures enforced by authorities.^3^ Although many countries have manged to control the initial spread of SARS-CoV-2 at the time of writing (beginning of May 2020), it is currently unclear how the pandemic will further develop and whether some countries will be affected by a second wave of sharply increasing COVID-19 case numbers in the foreseeable future.^4^ Hence, the current pandemic has been and may continue to be a challenge for health care systems and the medical workforce all around the world.

From research in physicians and nurses, it is well-known that work-related stressors such as working overtime are associated with impaired mental health, for example in the form of burnout, anxiety, and depression.^5–8^ Importantly, the consequences of reduced mental health not only affect HCWs themselves, but also the quality of care they provide and their professional functioning.^6,9–12^ This is highly problematic, because medical performance is essential to manage the consequences of public health crises. Therefore, monitoring and maintaining the mental health of HCWs is crucial during a pandemic. Moreover, a solid understanding of factors that influence HCWs’ mental health is needed to develop and optimize protective measures.^13^

Existing research on mental health of HCWs during a pandemic largely stems from the SARS outbreak at the beginning of this century.^14^ Several studies conducted in Canada have demonstrated the risk of pandemic-related stressors to HCWs’ mental health. For example, HCWs working in a clinical unit dedicated to the treatment of SARS patients experienced higher levels of stress than their peers.^15^ Another study found low organizational support as well as distrust in equipment to be associated with emotional exhaustion and anger.^16^ Moreover, a higher workload, assignment to unfamiliar tasks, health fears, and social isolation mediated the relationship between treating SARS patients and acute traumatic stress.^17^

Although these studies can provide preliminary evidence, their generalizability to the current SARS-CoV-2 pandemic is limited due to the peculiarities of each pandemic. Thus, timely research on mental health of HCWs during the current pandemic is needed. A first study was conducted in China by Lai et al. at the beginning of February 2020 among 1257 Chinese HCWs. These authors demonstrated that women, nurses, frontline workers, and those working in Wuhan, the epicenter of the pandemic, had elevated symptom levels of anxiety, depression, insomnia, and traumatic stress compared to men, physicians, second-line workers and those not working in Wuhan.^18^ In contrast, a second Chinese study conducted by Li et al. at the end of February 2020 found that 214 individuals from the general public and 292 non-frontline nurses reported higher levels of vicarious traumatization than 234 nurses working at the frontline.^19^ In a third Chinese study conducted between the end of February and the beginning of March, Zhang et al. reported that 927 medical HCWs had a higher prevalence of insomnia, anxiety, depression, somatization, and obsessive-compulsives symptoms than 1255 non-medical HCWs.^20^ Furthermore, being a woman was a significant predictor of insomnia, anxiety, and depression, and exposure to COVID-19 patients was a predictor of anxiety and insomnia. ^20^ However, a study among 470 HCWs in Singapore undertaken during the same period of time found the opposite, namely lower levels of stress-related symptoms in medical compared to non-medical HCWs.^21^ Lastly, in a qualitative study among 69 HCWs in the Unites States, Shanafelt et al. identified key concerns causing anxiety among HCWs including lack of access to appropriate personal protective equipment, support, and up-to-date information.^13^

Although these studies provide preliminary evidence for the mental health of HCWs during the current pandemic, their generalizability is limited in several ways. First, most studies were undertaken at the beginning of the pandemic, when the spread of the virus was mostly limited to a single province (in China) or to a few cases (in Singapore). Hence, sufficient equipment and manpower was still at hand or could be dispatched. Second, quantitative evidence was collected only in two countries, both located in Asia. Thus, differences in the experience with a pandemic outbreak of a respiratory virus (this experience is higher in Singapore and China compared to many other countries), differences in health care systems, and differences in cultural norms limit the generalization of the existing result to European or American countries. Third, although previous research explored a multiplicity of outcomes and associations, a comprehensive overview over these complex associations is lacking.

In a cross-sectional, nation-wide study, we assessed the mental health of physicians and nurses during the SARS-CoV-2 pandemic in Switzerland. The data was collected between March 28 and April 4, 2020, when the SARS-CoV-2 outbreak had reached the stage of a pandemic^1^. At that time, Switzerland had among the highest per capita rate of COVID-19 cases in the world. In addition to mental health data, based on the above-mentioned studies, we collected demographics (e.g., gender, profession, professional experience), work characteristics (e.g., availability of support, work hours), and data on COVID-19 exposure at work (e.g., exposure to COVID-19 patients, working as frontline staff) to investigate the influence of these variables on mental health.

The aims of our study were exploratory. The first aim was to assess HCWs’ mental health by their symptom levels of anxiety, depression, and burnout. Second, following previous studies, we aimed to compare levels of symptoms between subgroups (e.g., frontline and non-frontline workers). Finally, we conducted a network analysis to investigate the independent effect of the various factors outlined above on HCWs’ mental health. By this network analysis, we aimed to provide the first comprehensive overview of these factors and to visualize the interplay between them.

## Materials and Methods

This study had an explorative, cross-sectional design with a single period of data collection, and was carried out as a fully anonymous online survey. Inclusion criteria were defined as actively working as a nurse or physician in Switzerland. Given the explorative nature of the study design, no primary hypothesis was tested and thus no sample size calculation was conducted. Recruitment of participants was not aimed to be representative in a specific sense, but to ensure nation-wide participation. Thus, participants were recruited non-targeted through mailing lists of hospitals and professional societies, social media, and personal contacts of the study team members, with a focus of reaching health care workers in all parts of Switzerland. To reduce potential selection bias, the study was conducted in German, French and Italian.

The survey included questions regarding demographics, work characteristics, COVID-19 exposure, and mental health. T o account for the highly dynamic situation during the pandemic, the time period of reference for all questions of the survey was restricted to the past seven days. The survey was accessible through a link and could be filled out using a computer, tablet or smartphone. Data from participants was saved and thus accessible for analysis only after full completion of the survey. However, some items (e.g., years of professional experience) were assessed using a text-field, which lead to minor data loss due to wrong input by participants. The ethics committee of the canton Zurich assessed the study and officially declared that the study did not fall within the scope of the Human Research Act (BASEC-Nr. Req-2020-00471). Therefore, no authorization from the ethics committee was required.

Data was collected between March 28 and April 4, 2020, starting 2 weeks after the federal council (constituting the collective head of state) categorised the situation as “extraordinary” (March 16, 2020).^22^ With the declaration of this state of emergency, the federal council singed an executive order resulting in a partial “lock-down” (although this did not include a curfew).^22^

## Sample

We received a total of 1533 completed questionnaires. Of these, 120 (7.7%) participants did not meet the inclusion criteria of being a nurse or a physician. Of the remaining 1413 participants, 3 (0.2%) indicated their gender as “other” and were excluded from further analysis to ensure comparability of groups. This resulted in a final sample size of 1410, of which 857 (60.8%) were physicians and 553 (39.2%) were nurses.

## Measurements

### Demographics

Demographics included age (in years), gender (woman, man, and other), profession (physician, nurse, and other), professional experience (in years), and canton (corresponding to a federal state) in which participants worked.

### Work characteristics

Participants reported their average weekly work hours prior to the pandemic, their total work hours during the past seven days, and their average hours of sleep per night during the past seven days. Furthermore, using a Likert scale from 1 = “not at all” to 7 = “absolutely”, participants rated the extent to which they generally felt well equipped (e.g., with protective masks), well supported by the authorities and employers, and well informed (e.g., about the development of the pandemic) by the authorities and employers.

### COVID-19 exposure

Exposure to COVID-19 was assessed by several nominal questions (yes/no). First, participants indicated if they had experienced COVID-19 symptoms (e.g., fever, cough) since the beginning of the pandemic or if they had been tested positively for SARS-CoV-2. Second, they reported whether they had been exposed to suspected or confirmed COVID-19 patients during work, and third, whether they had been working in a clinical unit designated to diagnosis and treatment of COVID-19 patients. Participants answering to the latter question affirmatively were considered as frontline workers, the others as non-frontline workers.

### Mental health

The *General Anxiety Disorder-7* (GAD-7)^23^, a 7-item questionnaire, was used to measure symptoms of anxiety. Symptoms of depression were measured with the 9-item *Patient Health Questionnaire-9* (PHQ-9)^24^. Both questionnaires are validated and frequently used instruments to assess the self-reported symptom severity of generalized anxiety and depression.^25,26^ In both questionnaires, individual symptoms are assessed by ratings on a 4-point Likert scale ranging from 0 = “not at all” to 3 = “nearly every day”. An overall score can be calculated by summing individual items. Consequently, the sum scores of the GAD-7 and PHQ-9 range from 0 to 21 and 0 to 27, respectively. Sum scores of 10 points or higher indicate clinically relevant symptoms, corresponding to a diagnosis of generalized anxiety disorder or a depressive episodee.^23–25^ Burnout was assessed using a brief measurement tool for physician burnout developed and validated by West et al.^27^. This tool consists of two single items derived from the *Maslach Burnout Inventory* (MBI^28,29^) measuring emotional exhaustion and depersonalisation, two cardinal dimensions of burnout. These items were rated on a 7-point Likert scale ranging from 0 = “never” to 6 = “daily” and summed to form a total score. The answer format of all questionnaires was adapted to measure symptoms within the past seven days. The German, French, and Italian translations of the questionnaires provided by the corresponding manuals were used.

### Statistical analyses

Due to abnormally distributed data, continuous and ordinal items were described using the median and interquartile range [IRQ; 25%-75%] Categorical data was described with frequencies (%). Accordingly, we used two-tailed Mann-Whitney *U* tests and chi-square tests to assess differences between independent groups. Effect size of group differences of ordinal and continuous variables was assessed as rank biserial correlation. The significance level for all tests was set to alpha = .05. Given the explorative study design, *p* values were not adjusted for multiple comparisons. Descriptive statistics and comparison of independent groups were conducted using JASP version 0.11.^30^

To explore the complex relationships among demographic data, work characteristics, COVID-19 exposure, and symptoms of anxiety, depression, and burnout, we conducted a network analysis. To maximize power of the network analysis and due to intercorrelation among some of the assessed variables, we had to select items for the network analysis. Based on the previous studies on the current pandemic, we included gender, profession, exposure to COVID-19 patients, working as frontline staff, and perceived support by the employer. The latter factor was chosen from the variables assessing several aspects of support (i.e., feeling well equipped, well supported and well informed). Perceived support can be considered an umbrella term that, following traditional classifications of social support types,^31^ subsumes instrumental support (i.e., feeling well equipped) and informational support (i.e. feeling well informed). Furthermore, we included professional experience and work hours, since they are well-known risk factors for stress-related conditions such as burnout.^6,32^

Formally, the resulting network is a Gaussian Graphical Model, in which variables are represented by nodes, and edges between these variables represent partial correlations.^1^ Prior to network estimation, symptom overlap was tested using the default settings of the goldbricker function of the *networktools* package^33^. No exclusion of symptoms was suggested. The network was estimated using a regularization technique often used in psychopathology (e.g., Tibshirani^34^). The technique is based on the least absolute shrinkage and selection operator (LASSO^35,36^), which sets very small edges to zero and thus reduces the false positive rate. In other words, the technique is designed to have high specificity, while sensitivity might be limited (for more details see Epskamp et al.^37^). Stability and reliability analyses were conducted as recommended using the *bootnet* package.^37^ Network analysis was performed in the R statistical environment. Given that the network consisted of categorical, ordinal and continuous variables, the *mgm* package^38^ was used to estimate the network, and visualization was performed with the *qpgrah* package^39^.

## Results

### Overall Sample

Table 1 summarizes demographics, work characteristics, and COVID-19 exposure of the whole sample. Symptom severity scores of the whole sample are presented in Table 2. Of the finally included 1410 participants, the majority were German-speaking (n = 1124, 79.7%), women (n = 934, 66.2%), had a median age of 34 years [29-46] and median professional experience of 10 years [4-20]. Median working hours in the sample was 45 [36-54], with 572 (40.6%) participants working more hours than before the pandemic. Overall, experienced availability of medical equipment, support, and information by the employer and the authorities was high (all median scores ranging between 5 and 6, with 7 indicating the upper bound of the scale). One hundred ninety-seven (14%) of the participants had suspected COVID-19 symptoms or were tested positive for SARS-CoV-2, 1103 (78%) had contact with COVID-19 patients at work and 655 (46.5%) worked in designated COVID-19 units. Median anxiety and depressive symptom scores were 6 [3-10] and 5 [2-9]. Hence, these median scores were in the mild range (5 to 9 points^23,24^). Based on the suggested cut-offs (a total score of ≥ 10), 365 participants (25.9%) had clinically relevant symptoms of anxiety and 292 (20.7%) had clinically relevant symptoms of depression (see Table S2). On the 2-item burnout scale, scores ranged from 0 to 12, and the sample median was 4 [2-6].

**Table 1.** Demographics, Work Characteristics, and COVID-19 Exposure of 1410 Health Care Workers.

| Variable | Overall<br>(N = 1410) |  |
| --- | --- | --- |
|  | Median | IQR |
| **Demographics** |  |  |
| Age in years<sup>a</sup> | 34 | 29-46 |
| Women, n (%) | 934 | 66.2 |
| Professional experience in years<sup>b</sup> | 10 | 4-20 |
| Nurses, n (%) | 553 | 39.2 |
| Physicians, n (%) | 857 | 60.8 |
| German speaking, n (%) | 1124 | 79.7 |
| French speaking, n (%) | 143 | 10.1 |
| Italian speaking, n (%) | 143 | 10.1 |
| **Work Characteristics** |  |  |
| Total working hours in the previous 7 days | 45 | 36-54 |
| Total working hours per week prior to the pandemic<sup>c</sup> | 45 | 40-50 |
| Working more during the pandemic than before, n (%)<sup>c</sup> | 572 | 40.6 |
| Working less during the pandemic than before, n (%)<sup>c</sup> | 416 | 29.5 |
| Average number of sleep hours in the previous 7 days<sup>d</sup> | 7 | 6-7.5 |
| Having access to medical equipment | 5 | 3-6 |
| Perceived support by employer<sup>e</sup> | 6 | 4-7 |
| Perceived support by authorities | 5 | 3-6 |
| Perceived passage of information by employer<sup>f</sup> | 6 | 4-7 |
| Perceived passage of information by authorities | 5 | 4-6 |
| **COVID-19 Exposure** |  |  |
| Had suspected COVID-19 symptoms or tested positive for SARS-CoV-2, n (%) | 197 | 14.0 |
| Was exposed to suspected or confirmed COVID-19 patients at work, n (%) | 1103 | 78.2 |
| Worked in at clinical unit designated to diagnosis and treatment of patients with suspected or confirmed COVID-19, n (%) | 655 | 46.5 |

*Note.*<sup>a</sup> N = 1408; <sup>b</sup> N = 1384; <sup>c</sup> N = 1353; <sup>d</sup> N = 1334; <sup>e</sup> N = 1282; <sup>f</sup> N = 1290

**Table 2.** Mental Health of 1410 Health Care Workers and Comparison Across Different Subgroups.

|  |  | Gender |  |  | Profession |  |  | Workplace |  |  | Exposure to COVID-19 Patients |  |  |
| --- | --- | --- | --- | --- | --- | --- | --- | --- | --- | --- | --- | --- | --- |
|  | Overall<br>(N =1410) | Women<br>(n= 934) | Men<br>(n= 476) | Effect<br>size | Nurses<br>(n= 553) | Physicians<br>(n= 857) | Effect<br>size | Frontline<br>(n= 655) | Secondline<br>(n= 755) | Effect<br>size | Yes<br>(n= 1103) | No<br>(n= 307) | Effect<br>size |
| Anxiety | 6 (3-10) | 6 (4-10.75) | 4 (3-8)*** | -.197 | 7 (4-10) | 5 (3-9)*** | -.144 | 6 (3-11) | 5 (3-9)** | -.095 | 6 (3-10) | 5 (3-8)*** | -.157 |
| Depression | 5 (2-9) | 6 (3-9) | 4 (2-7)*** | -.167 | 6 (3-9) | 5 (2-8)*** | -.142 | 6 (3-10) | 4 (2-8)*** | -.145 | 6 (3-9) | 4 (1-7)*** | -.218 |
| Burnout | 4 (2-6) | 4 (2-6) | 4 (2-6) |  | 4 (2-7) | 4 (2-6)* | -.076 | 5 (2-7) | 3 (2-6)*** | -.200 | 4 (2-7) | 3 (1-5)*** | -.251 |

*Note.*
ES = Effect Size; Effect size is measured as a rank-biserial-correlation; Burnout = Overall burnout symptom score; Anxiety = Overall GAD-7 score; Depression = Overall PHQ-9 score.
\* p < 0.05, \*\* p < 0.01, \*\*\* p < 0.001

### Group differences

Results from group comparisons of symptom severity are presented in Table 2. In summary, women had higher symptom levels of anxiety and depression than men, yet similar burnout symptoms as men. Nurses, frontline staff and HCWs exposed to COVID-19 patients showed more symptoms of anxiety, depression, and burnout than physicians, non-frontline staff and non-exposed HCWs. However, all effects of the group comparisons were small (ranging from −.076 to −.251). Similar to these results, a significantly higher share of women, nurses, frontline staff and HCWs exposed to COVID-19 patients had clinically relevant symptoms of anxiety and depression compared to their male, physician, non-frontline and non-exposed colleagues (see Table S2). Differences between groups with regard to demographics and additional variables are presented in Table S1.

### Relationships among the investigated variables (network analysis)

The results of the network analysis are visualized in Figure 1. The edges in the network represent partial correlations between the variables, with the thickness of the edge representing the magnitude of the correlation and the colour indicating the direction (red = negative, blue = positive). Expectedly, being a woman was associated with working as a nurse, and working in a designated COVID-19 unit was associated with exposure to COVID-19 patients at work. The total symptom scores of anxiety, depression, and burnout were associated with one another. Symptoms of depression were not associated with any factor other than burnout and anxiety. Regarding anxiety, associations with gender, professional experience and perceived support by the employer emerged. Burnout was associated with professional experience, work hours, exposure to COVID-19 patients, and perceived support by the employer.

**Figure 1.**
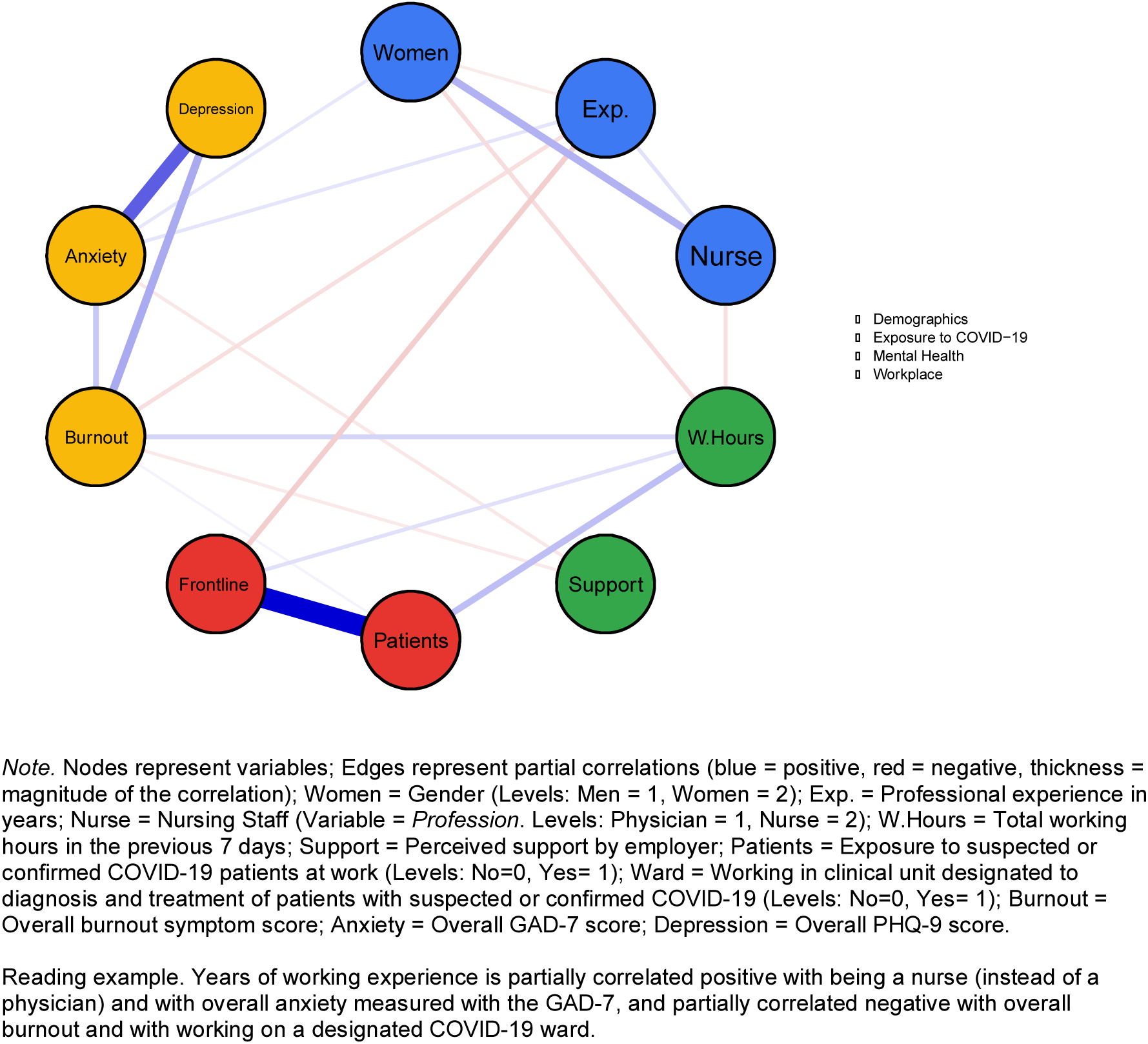
Relationships between demographics, work characteristics, COVID-19 exposure, and symptoms of anxiety, depression, and burnout.

## Discussion

This study represents the first nation-wide survey on mental health and work-related strain in health care workers during the SARS-CoV-2 pandemic in a Western country. Overall, approximately 40% of our sample worked more during the pandemic than before. Almost half of the sample was assigned to a designated COVID-19 unit and close to 80% were exposed to suspected or confirmed COVID-19 patients at work. Importantly, health care workers felt mostly well equipped, supported and informed by the employers and the authorities.

In general, participants reported mild levels of anxiety and depression, and elevated burnout scores. Still, overall anxiety and depression were significantly higher in our sample than the study by Lai et al. who investigated a sample of Chinese HCWs with the same questionnaires and also higher than a in Singaporean HCWs^21^. Compared to the Chinese nurses assessed by Zhang et al.^20^, nurses in our sample had similar levels of anxiety and depression. The higher prevalence of anxiety and depression among Swiss HCWs could have several reasons. First, at the time of data collection, Switzerland had very high per capita rate of COVID-19 cases, higher than during the study period of the other two studies. Second, in non-pandemic times, several studies have consistently documented higher levels of anxiety and depression in European countries compared to Asian nations.^40^ Thus, the higher prevalence in our sample might be due to higher general levels of anxiety and depression or to culture-dependent, social desirability effects. Third, in contrast to Singapore and China, Switzerland was not affected by the SARS pandemic at the beginning of the century. Thus, Chinese and Singaporean HCWs probably had more experience dealing with a pandemic than Swiss HCWs, which could have reduced their symptom burden.^41^

Women, nurses, frontline staff, and health care workers exposed to COVID-19 patients exhibited higher levels of symptoms and a higher prevalence of clinically relevant symptoms of anxiety and depression when compared to their male, physician, non-frontline and non-exposed peers. However, all effects were small. The found differences between women and men, nurses and physicians, and frontline and non-frontline HCWs are in accordance with the study of Lai et al.^18^, thus replicating their results. In contrast, Li et al. reported that vicarious, i.e. indirect forms of traumatization was lower among HCWs working at the frontline compared to other HCWs and the general public.^19^ Still, the comparability of their results is limited, as they investigated a different aspect of psychopathology, namely vicarious traumatization, than Lai et al. and we did. However, unadjusted comparisons across several groups must be interpreted with caution, because some variables (e.g., women and nurse or frontline workers and exposure to COVID-19 patients) are highly intercorrelated and thus confound results.

By conducting the first network analysis on mental health and associated factors during the pandemic, we were able to map relationships between several variables while controlling for their mutual influence. Here, we highlight some specific associations. While support by the employer was associated with anxiety and burnout, working in a COVID-19 ward was not. Exposure to COVID-19 patients was only associated with burnout. Hence, although we found group differences in mental health between frontline and non-frontline staff and between HCWs who have and have not been exposed to COVID-19 patients, these associations did not remain significant after adjusting for all factors within the network. Taken together, COVID-19 exposure only partially predicted burnout, while support by the employer was a significant predictor of both burnout and anxiety. The role of perceived support has been studied extensively in occupational and health psychology,^31^ and its direct relationship with mental health of HCWs is well documented.^6,7,42^ For example, as postulated by the job-demand-control-support model^43^, which has a broad empirical foundation,^44^ support is not only important to well-being but reduces the mental strain caused by job demands such as, in this case, COVID-19 exposure. Moreover, the importance of support was also recently emphasized in a qualitative study on HCWs’ concerns with regard to the SARS-CoV-2 pandemic.^13^ Another possible interpretation for the absence of individual effects of COVID-19 variables is that these effects were very small and thus did not survive regularization. In this case, even if the effects exist, they would not be of a clinically relevant magnitude.

Regarding anxiety, associations with being a woman and higher professional experience in addition to perceived support by the employer emerged. While higher levels of anxiety among women in general^45^ and in female HCWs during a pandemic in particular^18,20^ are well documented, the positive association with professional experience is counterintuitive. Given that HCWs with a higher professional experience tended to be older, we can only speculate that these HCWs more likely belonged to a risk group for COVID-19 related complications (e.g., cardiac diseases). This might have led to more anxiety. Burnout was negatively associated with professional experience and positively related to work hours, which is well-documented in literature on HCW burnout.^6,32^

This study is limited in several ways. First, the cross-sectional nature of the study with a single period of data collection and no control group does not allow to draw conclusions about changes in symptoms. In other words, we do not know whether symptoms changed compared to before the pandemic, whether changes are a consequence of the pandemic, or whether HCWs reacted in a specific way different from the general population. These important questions need to be addressed by future studies with an appropriate design (e.g., within the frame of ongoing cohort studies). Second, given the non-targeted recruitment, our sample was likely not representative of HCWs in Switzerland. Moreover, the non-targeted recruitment might have introduced several kinds of selection bias (e.g., very busy HCWs might not have been willing to participate). In addition, due to the non-targeted recruitment, we are unable to calculate a response rate. Still, the 857 physicians participating in this study represent approximately 2.3% of all 37’882 licenced Swiss physicians in 2019.^46^ This exceeds the per capita rate of participating HCWs of previous studies by far. Third, mental health of participants was measured using self-report questionnaires. This might lead to an overestimation of symptoms.^24^ Fourth, the adaptation of all questionnaires to cover symptom experience over the last seven days has not been validated and limits comparability to studies undertaken with the original validated versions of the questionnaires (covering two weeks in case of the GAD-7^23^ and PHQ-9^24^) and a full year in case of the brief measurement tool for physician burnout developed and validated by West et al.^27^). However, the strength of the restriction to the past seven days lies in the capacity to measure symptoms during a highly dynamic time of crisis. Finally, questions regarding COVID-exposure or perceived support were developed for this specific study and were therefore not validated.

Notwithstanding these limitations, our study has clinical and scientific implications. Our finding that 25.9% of the investigated HCWs had clinically relevant symptoms of anxiety and 20.7% clinically relevant symptoms of depression underscores the importance to systematically monitor HCWs’ mental health during this ongoing pandemic. Furthermore, given that perceived support and higher levels of anxiety and burnout were negatively related, we encourage the implementation of supportive measures for HCWs’ well-being during this crisis. Such measures should address key concerns of HCWs identified in previous research (e.g., sufficient access to personal protective equipment and access to child-care during increased work hours^13^). Most importantly, however, HCWs themselves can best express their individual needs. Hence, besides a systematic monitoring of HCWs’ mental health, we encourage managers and regulators to actively engage with the health care force and hear them and their concerns. Due to the well-documented negative effect of impaired mental health of HCWs on their provided care,^6,9–12^ these measures not only support HCWs themselves but also serve patients by ensuring continuation of high-quality care, especially during a public health crisis.

Regarding future research, our study implies the need to address remaining questions with adequately designed studies. For example, changes in symptoms during different stages of the pandemic should be addressed by longitudinal studies. In addition, existing cohort studies should answer the question whether HCWs developed more symptoms during the pandemic compared to before.

In conclusion, in our sample, of which 40% worked more hours than prior to the pandemic, overall symptom levels of anxiety and depression were mild, and burnout was elevated. Still, symptoms of anxiety and depression were significantly higher than in a similar study conducted in China during the beginning of the SARS-CoV-2 pandemic.^18^ In general, participants felt well-equipped and well-supported by their employer and the authorities. Women reported more symptoms than men, nurses more than physicians, frontline staff more than those not working at the frontline, and HCWs exposed to COVID-19 more than non-exposed peers. However, these effects were all small and most of them did not remain significant after controlling for the other factors within the network analysis. Importantly, whereas COVID-19 exposure was only partially associated with burnout, perceived support by the employer independently predicted anxiety and burnout. Given that the pandemic is ongoing and its future progress unpredictable, we encourage the implementation of monitoring systems for HCWs’ mental health and measures to maintain their well-being during this crisis.

## Data Availability

Data is available on reasonable request for non-commercial use.

## Acknowledgments

We would like to express our gratitude to all professional societies and hospitals who supported us and kindly distributed our survey among health care workers. Our special thanks go to Sophie-Charlotte Fischer.

## Address for correspondence

Tobias R. Spiller, MD., Department of Consultation-Liaison Psychiatry and Psychosomatic Medicine, University Hospital Zurich, Haldenbachstrasse 16/18, CH-8091, Zurich, Switzerland;

## Supplementary Materials for

**Table S1.** Demographics, Work Characteristics, and COVID-19 Exposure Across Different Subgroups

|  | Overall<br>(N = 1410) | Gender |  |  |
| --- | --- | --- | --- | --- |
|  |  | Women<br>(n= 934) | Men<br>(n= 476) | Effect size |
| **Demographics** |  |  |  |  |
| Age in years median (IQR)<sup>a</sup> | 34 (29-46) | 34 (29-45) | 35 (30-47)** | .085 |
| Women, n (%) | 476 (33.8) |  |  |  |
| Professional experience in years, median (IQR)<sup>b</sup> | 10 (4-20) | 10 (4-20) | 9 (3-20) |  |
| **Work characteristics** |  |  |  |  |
| Total working hours in the previous 7 days, median (IQR)<sup>c</sup> | 45 (36-54) | 43 (34-50) | 50 (42-58.5)*** | .256 |
| Total working hours per week prior to the pandemic, median (IQR)<sup>d</sup> | 45 (40-50) | 42 (33.3-50) | 50 (43-55)*** | .418 |
| Working more during the pandemic than before, n (%)<sup>d</sup> | 572 (40.6) | 387 (41.3) | 185 (38.9) |  |
| Working less during the pandemic than before, n (%)<sup>d</sup> | 416 (29.5) | 254 (27.2) | 162 (34.0)* |  |
| Average number of sleep hours in the previous 7 days, median (IQR)<sup>e</sup> | 7 (6-7.5) | 7 (6-7.5) | 7 (6-7) |  |
| Having access to medical equipment, median (IQR) | 5 (3-6) | 5 (3-6) | 5 (3-6)** | .087 |
| Perceived support by employer, median (IQR)<sup>f</sup> | 6 (4-7) | 6 (4-7) | 6 (4-6) |  |
| Perceived support by authorities, median (IQR) | 5 (3-6) | 5 (3-6) | 5 (3-6) |  |
| Perceived passage of information by employer, median (IQR)<sup>g</sup> | 6 (4-7) | 6 (4-7) | 6 (5-7) |  |
| Perceived passage of information by authorities, median (IQR) | 5 (4-6) | 6 (4-6) | 5 (4-6) |  |
| **COVID-19 Exposure** |  |  |  |  |
| Had suspected COVID-19 symptoms or tested positive for SARS-CoV-2, n (%) | 197 (14.0) | 125 (13.4) | 72 (15.1) |  |
| Was exposed to suspected or confirmed COVID-19 patients at work, n (%) | 1103 (78.2) | 712 (76.2) | 391 (82.1)** |  |
| Worked in at clinical unit designated to diagnosis and treatment of patients with suspected or confirmed COVID-19, n (%) | 655 (46.5) | 412 (45.1) | 234 (49.2) |  |

**Table S1 (continued)**
|  | Profession |  |  | Workplace |  |  | Exposure to COVID-19 Patients |  |  |
| --- | --- | --- | --- | --- | --- | --- | --- | --- | --- |
|  | Nurses<br>(n= 553) | Physicians<br>(n= 857) | Effect size | Frontline<br>(n= 655) | Secondline<br>(n= 755) | Effect size | Yes<br>(n= 1103) | No<br>(n= 307) | Effect size |
| **Demographics** |  |  |  |  |  |  |  |  |  |
| Age in years, median (IQR)<sup>a</sup> | 37 (29-48) | 34 (29-45) |  | 32 (29-41) | 38 (30-50.75)*** | .260 | 33 (29-44) | 41 (31-52)*** | .224 |
| Women, n (%) | 477 (86) | 457 (53.3)*** |  | 421 (64.3) | 513 (67.9) |  | 712 (64.6) | 222 (72.3)* |  |
| Professional experience in years, median (IQR)<sup>b</sup> | 14 (7-25) | 7 (3-18)*** | -.284 | 7 (3-15) | 14 (5-25)*** | .265 | 8 (3.5-19) | 16 (6 -26)*** | -.241 |
| **Work characteristics** |  |  |  |  |  |  |  |  |  |
| Total working hours in the previous 7 days, median (IQR)<sup>c</sup> | 42 (34-50) | 50 (40-55)*** | .227 | 50 (40-57) | 42.5 (30-50)*** | -.259 | 48 (40-55) | 40 (25-48)*** | -.367 |
| Total working hours per week prior to the pandemic, median (IQR)<sup>d</sup> | 40 (30-42) | 50 (45-55)*** | .687 | 48 (40-50) | 42 (34-50)*** | -.161 | 45 (40-50) | 42 (30-50)*** | -.224 |
| Working more during the pandemic than before, n (%)<sup>d</sup> | 309 (58.5) | 263 (31.9)*** |  | 304 (46.4) | 268 (35.5)*** |  | 490 (46.4) | 82 (27.7)*** |  |
| Working less during the pandemic than before, n (%)<sup>d</sup> | 71 (13.4) | 345 (41.8)*** |  | 163 (24.9) | 253 (33.5)*** |  | 298 (28.2) | 118 (39.9)*** |  |
| Average number of sleep hours in the previous 7 days, median (IQR)<sup>e</sup> | 6 (6-7) | 7 (6-7.5)*** | .143 | 7 (6-7.5) | 7 (6-7.5) |  | 7 (6-7) | 7 (6-8) |  |
| Having access to medical equipment, median (IQR) | 4 (3-6) | 5 (3-6)*** | .128 | 5 (3-6) | 5 (3-6) |  | 5 (3-6) | 5 (3-6) |  |
| Perceived support by employer, median (IQR) | 5 (4-6) | 6 (4-7)*** | .115 | 6 (4-6) | 6 (4-7) |  | 5 (4-6) | 6 (5-7)* | -.092 |
| Perceived support by authorities, median (IQR) | 5 (3-6) | 5 (3-6)*** | .116 | 5 (3-6) | 5 (3-6) |  | 5 (3-6) | 5 (3.5-6)** | -.112 |
| Perceived passage of information by employer, median (IQR) | 6 (4-7) | 6 (5-7)* | .079 | 6 (4-7) | 6 (5-7) |  | 6 (4-7) | 6 (5-7)** | -.106 |
| Perceived passage of information by authorities, median (IQR) | 5 (4-6) | 6 (4-6) |  | 5 (4-6) | 6 (4-6) |  | 5 (4-6) | 6 (5-6.5)* | -.090 |
| **COVID-19 Exposure** |  |  |  |  |  |  |  |  |  |
| Had suspected COVID-19 symptoms or tested positive for SARS-CoV-2, n (%) | 76 (13.7) | 121 (14.1) |  | 89 (13.6) | 108 (14.3) |  | 162 (14.7) | 35 (11.4) |  |
| Was exposed to suspected or confirmed COVID-19 patients at work, n (%) | 416 (75.2) | 688 (80.2)* |  | 627 (95.7) | 476 (63.0)*** |  |  |  |  |
| Worked in at clinical unit designated to diagnosis and treatment of patients with suspected or confirmed COVID-19, n (%) | 240 (43.3) | 415 (48.4) |  |  |  |  | 627 (56.8) | 28 (9.1)*** |  |

**Table S2.**
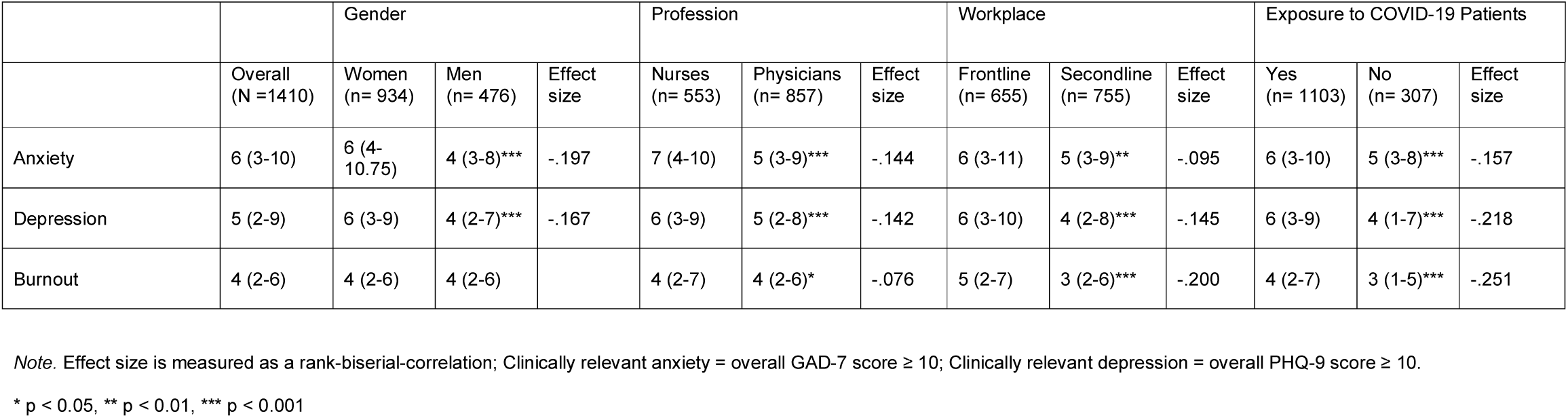
Clinically Relevant Symptoms of Anxiety and Depression of 1410 Health Care Workers Across Different Subgroups

**Figure S1.**
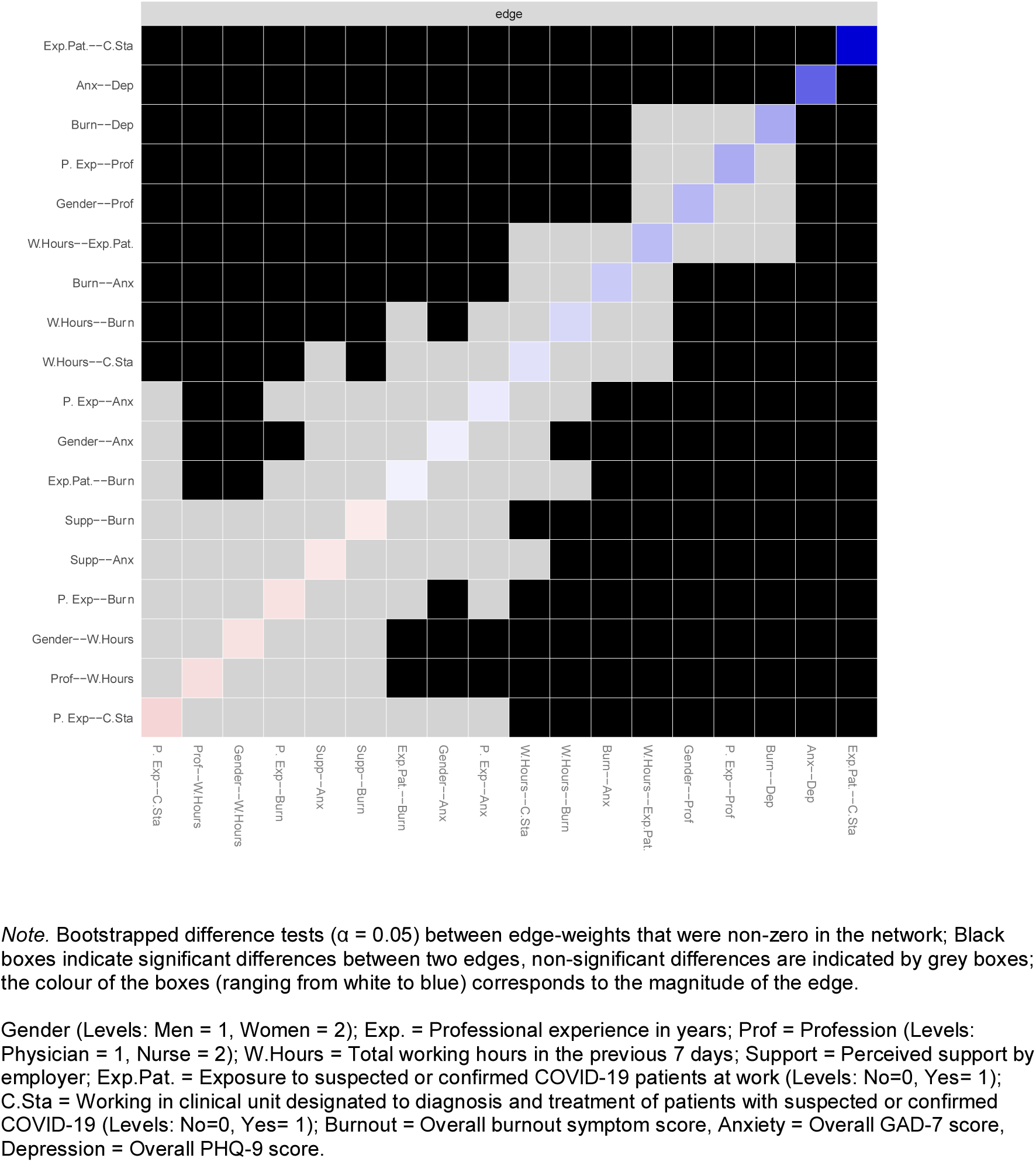
Bootstrap edge weights difference test between non-zero estimated edge-weights in the network shown in Figure 1.

**Figure S2.**
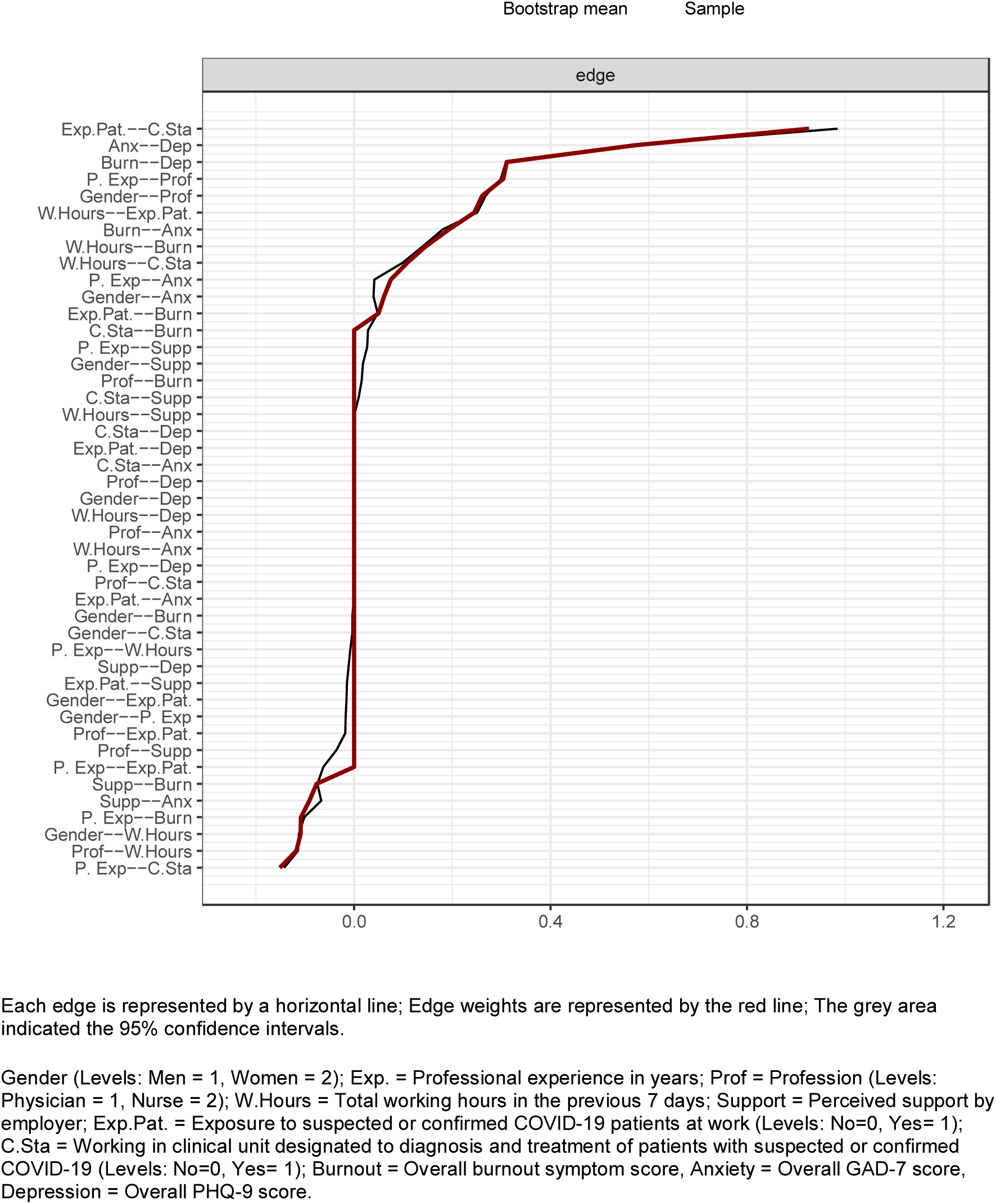
Bootstrap 95% confidence intervals for estimated edge weights of the network.

## Footnotes

1 Conceptually, the partial correlations can be understood as regression coefficients resulting from repeated regression analyses. In each individual regression analysis, a given node is the outcome (dependent variable) and all remaining nodes are the predictors (independent variables). This is repeated for all nodes. However, this is not the way the analysis is conducted (see above).

